# Proteomic profiling of the large vessel vasculitis spectrum identifies shared signatures of innate immune activation and stromal remodelling

**DOI:** 10.1101/2024.09.06.24313218

**Authors:** Robert T. Maughan, Erin MacDonald-Dunlop, Lubna Haroon-Rashid, Louise Sorensen, Natalie Chaddock, Shauna Masters, Andrew Porter, Marta Peverelli, Charis Pericleous, Andrew Hutchings, James Robinson, Taryn Youngstein, Raashid A. Luqmani, Justin C. Mason, Ann W. Morgan, James E. Peters

## Abstract

Takayasu arteritis (TAK) and giant cell arteritis (GCA) are the primary forms of large vessel vasculitis (LVV) and can result in serious cardiovascular morbidity. Improved understanding of the molecular basis of these diseases is required to develop novel biomarkers and targeted treatments. Moreover, it is unclear whether shared or distinct pathogenic processes underpin the LVV spectrum. To address this, we performed plasma proteomic profiling, quantifying 184 plasma proteins using Olink immunoassays in two independent cohorts totalling 405 individuals. In Cohort 1, comparison of patients with TAK (N=96) and large vessel-GCA (LV-GCA) (N=35) versus healthy controls (HCs) (N=35) revealed 52 and 72 significant differentially abundant proteins, respectively. Correlation with disease activity status identified novel TAK and LV-GCA disease activity markers. Cohort 2 consisted of patients presenting acutely with possible cranial GCA (C-GCA); C-GCA was subsequently confirmed (n=150) or excluded (n=89). 31 proteins were associated with C-GCA. Analyses stratified by temporal artery biopsy results revealed enrichment of the proteomic signal in biopsy-proven GCA, suggesting the presence of distinct endotypes within C-GCA. Cross-disease comparison revealed that active TAK, LV-GCA and biopsy-proven C-GCA had highly similar plasma proteomic profiles. Twenty-six proteomic associations were shared across all three groups including IL6, monocyte/macrophage related proteins (CCL5, CCL7, CSF1), tissue remodelling proteins (VEGFA, TIMP1, TNC) and proteins not previously linked to LVV (TNFSF14, IL7R). We also observed disease-specific associations including increased CXCL9 in LV-GCA and C-GCA but not in TAK and decreases in the extracellular matrix protein COMP in TAK but not in LV-GCA or C-GCA. Evaluation of publicly available transcriptomic data from LV-GCA aortic tissue revealed that 47 of the 112 proteins significantly altered in ≥1 LVV type had significantly altered mRNA expression in LVV aortic tissue. Similarities in LVV proteomic profiles suggest shared pathobiology involving innate immunity, particularly monocyte/macrophages, lymphocyte homeostasis and tissue remodelling processes. Our results highlight a signature of immune-stromal cross talk in LVV and identify potential novel therapeutic targets in this axis (e.g. TNFSF14). The correspondence of plasma signatures to tissue phenotype highlights the potential for non-invasive monitoring of arterial inflammation and injury.

## Introduction

Takayasu arteritis (TAK) and giant cell arteritis (GCA), the most common forms of large-vessel vasculitis (LVV) in adults, are characterised by granulomatous arterial inflammation. Progressive damage to arterial walls typically results in stenotic remodelling with consequent tissue ischaemia and manifestations such as sight loss, stroke, myocardial infarction and limb claudication^1^. Despite phenotypic similarities, TAK and GCA have different demographics, particularly age of onset, and, to a lesser extent, they affect different arterial territories. TAK affects the aorta and its major branches, while classically GCA has been described as involving the cranial arteries (C-GCA) such as the temporal artery. However, following advances in non-invasive vascular imaging techniques, it became clear that the aorta and other large vessels are frequently affected in GCA, and some patients have a large vessel-type presentation (LV-GCA) with non-specific constitutional symptoms and/or limb claudication similar to TAK^2,3^. Frequent large-vessel involvement in GCA and similar histopathological changes has led to debate regarding whether TAK and GCA represent varying manifestations of the same disease^4,5^. This question has important implications for drug development and clinical trial design. However, such comparisons are currently limited by an incomplete understanding of the pathogenic underpinnings of these diseases and a lack of comparative molecular data across LVV phenotypes.

There are several important challenges in the clinical management of TAK and GCA. Both initial diagnosis and recognition of relapse may be delayed and are made more difficult by a lack of effective biomarkers^6^. Blood tests such as C-reactive protein (CRP) lack specificity while vascular imaging can be insensitive, particularly in glucocorticoid-treated patients, and impractical for frequent serial monitoring^7,8^. In the era before widespread access to ultrasound scans, the diagnosis of C-GCA was confirmed by performing a temporal artery biopsy. While considered the “gold standard” diagnostic test, the presence of skip lesions may lead to false negative results. A negative biopsy therefore does not exclude the diagnosis of GCA. Progress in the treatment of LVV, particularly the development of targeted biologic therapy, lags behind that of other rheumatic diseases. Accordingly, there is overreliance on long-term glucocorticoids to maintain disease control with resulting iatrogenic harm^9,10^. Thus, there is a need for better biomarkers and novel therapeutics to improve patient outcomes.

Proteomic profiling has the potential to address these challenges^11^. Proteins are the effector molecules of most biological functions and the targets of most drugs. Given the proximity of arterial tissue to the circulation, blood-based proteomics is likely to be informative in LVV. Specifically, we hypothesised that the levels of inflammation- and cardiovascular-related proteins will provide a read-out of disease activity and arterial pathobiology in LVV patients and enable evaluation of molecular similarities and differences between GCA and TAK. To this end, we performed proteomic profiling of 184 circulating proteins in two independent cohorts that included 281 patients with TAK, LV-GCA or C-GCA. We identify protein signatures associated with each LVV type and with disease activity. Cross-disease comparison revealed a striking similarity between the proteomic profiles of active LVV types. Our data indicate the shared dysregulation of innate immune and tissue remodelling pathways and highlight the potential for therapeutics targeting immune-stromal cross-talk.

## Results

To identify proteins associated with LVV and to evaluate the presence of shared or distinct molecular signatures across TAK, LV-GCA and C-GCA, we measured plasma levels of 184 inflammation- and cardiovascular-related proteins (**Table S1**) in two independent cohorts using the Olink Target antibody-based proximity extension assay (PEA) (**Figure 1**). To provide a standardised nomenclature, we report proteins using the non-italicised HUGO gene symbol of the encoding gene.

**Figure 1.**
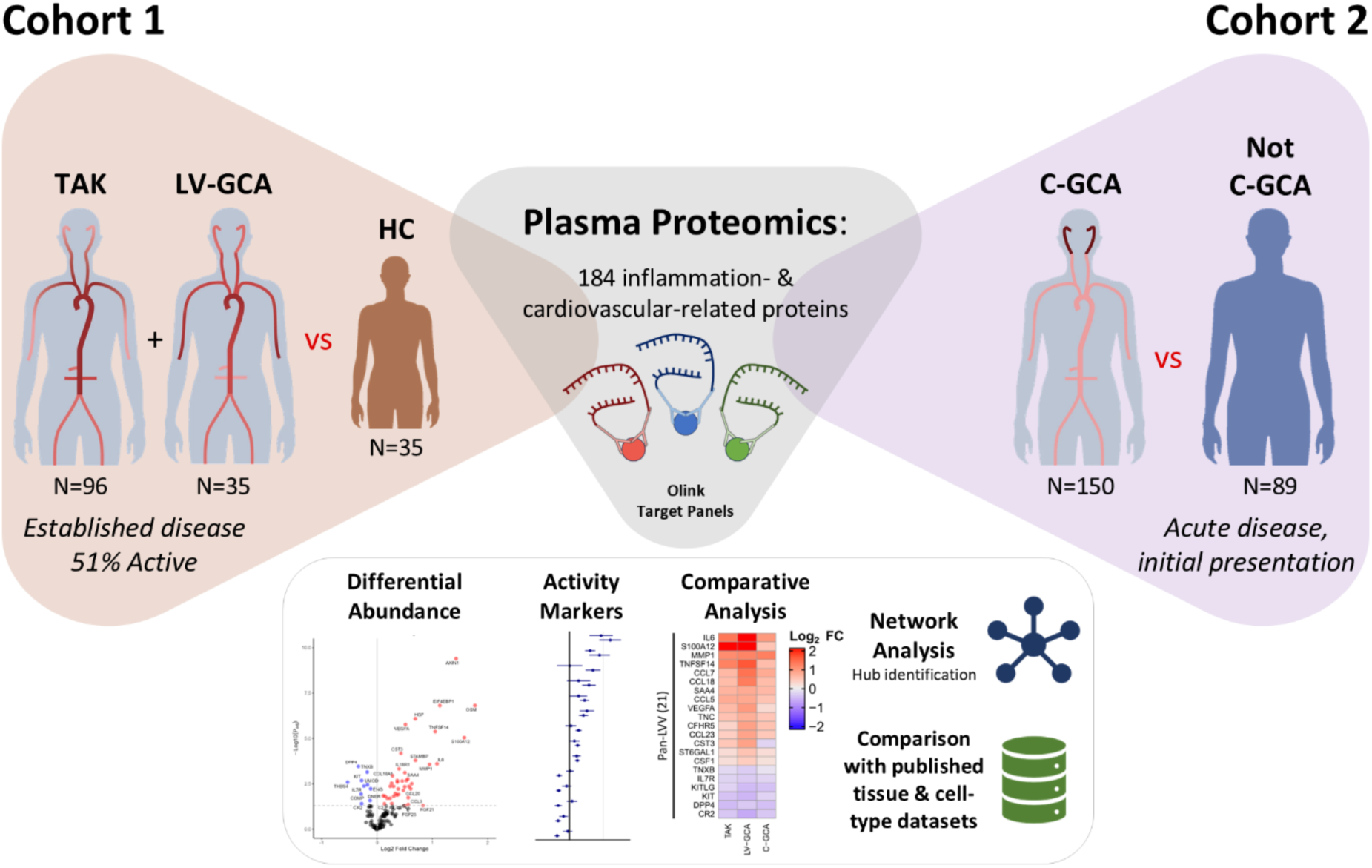
Study Overview. Schematic overview of study to investigate the plasma proteomic changes associated with each form of large vessel vasculitis in two independent cohorts. Cohort 1: patients with established Takayasu arteritis (TAK) and large vessel giant cell arteritis (LV-GCA) and healthy control (HC) participants. Cohort 2: patients presenting with possible cranial giant cell arteritis who subsequently had diagnosis confirmed (C-GCA) or ruled out (Not C-GCA). Disease-specific proteomic profiles defined by differential abundance analysis were compared. Network-based analysis of LVV-associated proteins and integrated analysis with published tissue and cell-type datasets was also conducted.

### Shared plasma proteomic profiles in TAK and LV-GCA

Cohort 1 included 96 TAK patients, 35 LV-GCA patients and 35 healthy controls (HCs) (**Table 1**). Patient characteristics were typical for TAK and LV-GCA with regards to age and sex, with younger onset and greater female:male ratio in TAK. HCs were well matched in terms of demographics to the TAK patients. LV-GCA patients were older and had a higher proportion of individuals of White European ancestry. The patient samples analysed encompassed a broad range of disease activit.0y, disease duration and treatment status, particularly in TAK where sample size was greater. Patients with active disease tended to have shorter disease durations and were receiving higher glucocorticoid doses, as might be expected (**Table S2**). In Cohort 1, 158 proteins (86%) passed quality control (QC) parameters (**Methods**) and were available for analysis.

**Table 1:** TAK and LV-GCA cohorts.

|  | HC | TAK | LV-GCA |
| --- | --- | --- | --- |
| N | 35 | 96 | 35 |
| Female (%) | 31 (88.6) | 91 (94.8) | 28 (80) |
| Age (years) | 38.2 [30.4, 52.5] | 41.6 [30.9, 55.6] | 67.2 [61.3, 72.3] |
| Ethnicity |  |  |  |
| White European (%) | 21 (60) | 55 (57.3) | 30 (85.7) |
| Asian (%) | 11 (31.4) | 35 (36.5) | 4 (11.4) |
| Other (%) | 3 (8.6) | 6 (6.3) | 1 (2.9) |
| Time since diagnosis, yrs | - | 3.9 [1.1, 10.3] | 1.4 [0.5, 3.7] |
| Active Disease (%)* | - | 56 (58.3) | 11 (31.4) |
| CRP >5mg/L (%) | - | 42 (43.7) | 17 (48.6) |
| ESR >20mm/hr (%) | - | 59 (61.5) | 17 (48.6) |
| Treatment |  |  |  |
| No treatment (%) | - | 21 (21.9) | 6 (17.1) |
| Time on Treatment | - | 2.6 [0.7, 7.4] | 1.4 [0.5, 4.1] |
| Glucocorticoids (%) | - | 62 (64.6) | 28 (80) |
| csDMARD (%) | - | 57 (59.4) | 18 (51.4) |
| Biologic (%) | - | 7 (7.3) | 1 (2.8) |

Data is median [IQR] or N (%) where indicated. \*For Takayasu Arteritis patients (TAK), active disease: ITAS2010 score $\geq 1$ or ITAS.CRP $\geq 2$ ; for large vessel giant cell arteritis patients (LV-GCA): NIH score $\geq 2$ at the time of sampling. CRP, C-reactive protein; ESR, erythrocyte sedimentation rate. Treatment at time of sampling is summarised. csDMARD, conventional synthetic disease modifying anti-rheumatic drug; Biologic, targeted biologic agent e.g. anti-IL6R monoclonal antibodies or TNF inhibitor.

Comparison of proteomic profiles between TAK patients and HCs identified 52 differentially abundant proteins (DAP), with 42 upregulated and 10 downregulated (**Figure 2A, Table S3**). Cross-referencing of our results to those of a previous study^12^ which used a different proteomic platform demonstrated that many of our proteomic associations were novel (**Figure S1**, **Table S3**). We next compared LV-GCA to HCs, revealing 72 DAP, with 60 increased and 12 decreased (**Figure 2B, Table S4**). The proteomic changes in TAK and LV-GCA in comparison to HCs were highly similar; 40 proteins were significantly altered in both diseases and 85% of all proteins had directionally concordant changes compared to HCs (**Figure 2C**). Proteomic similarity of TAK and LV-GCA was also reflected in principal component analysis (PCA), with TAK and LV-GCA clustering together and separated from HCs (**Figure S2A**).

**Figure 2.**
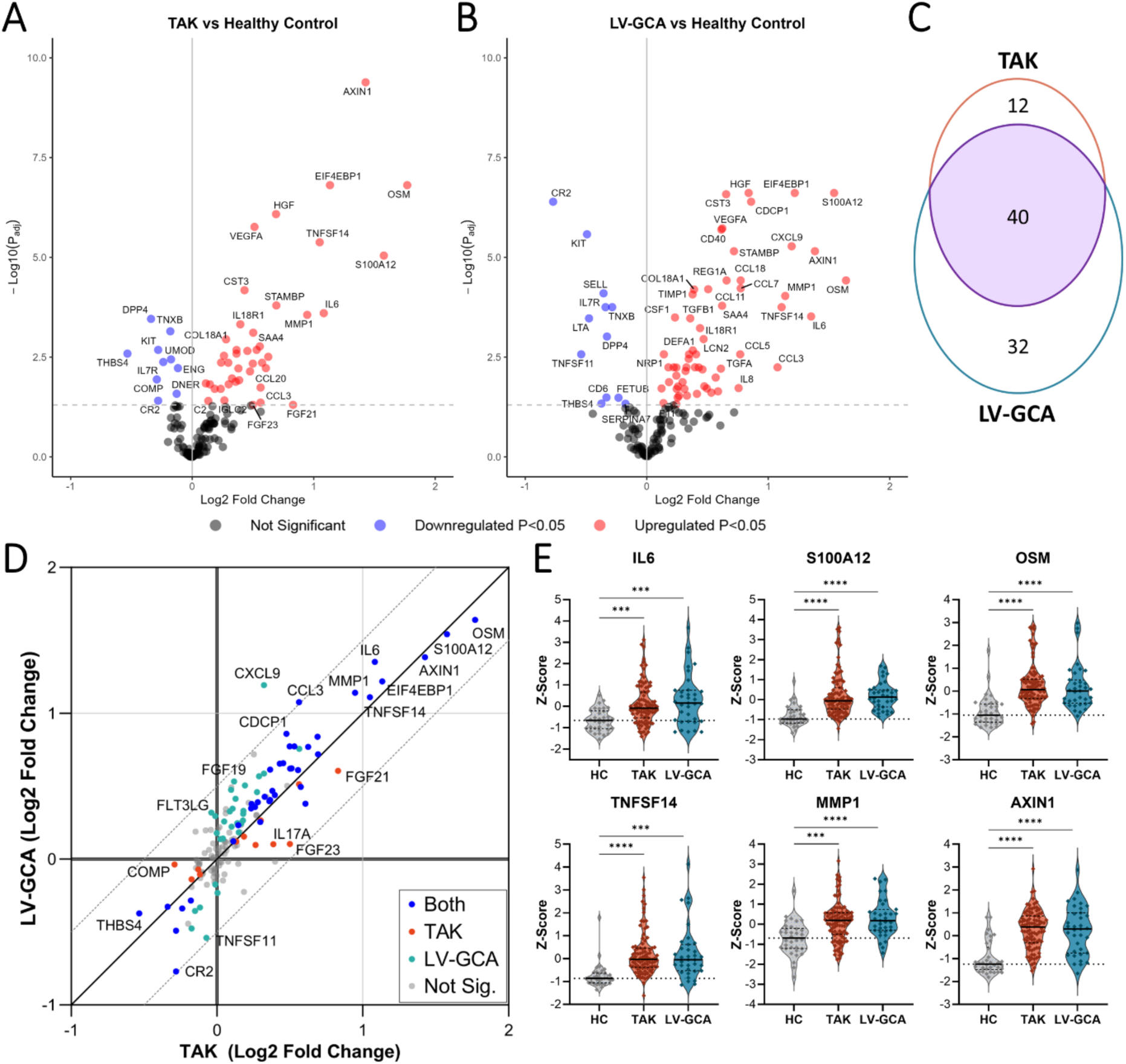
Plasma proteomic changes in TAK and LV-GCA compared to Healthy Control participants. Volcano plots showing results of differential protein abundance analyses: A) Takayasu Arteritis patients (TAK, N=96) vs healthy controls (HC, N=35) B) Large Vessel Giant Cell Arteritis (LV-GCA, N=35) vs HC. −Log10(P_adj_) = −log_10_ Benjamini-Hochberg adjusted p-value. Red and blue indicate proteins that are significantly (P_adj_ < 0.05) upregulated and downregulated, respectively. C) Venn diagram showing the overlap in proteins significantly altered in TAK vs HC compared to LV-GCA vs HC analyses. D) Comparison of log_2_ fold changes in all proteins for TAK vs HC and LV-GCA vs HC analyses, diagonal lines represent line of identity and +/- 0.5 log_2_ fold change. Blue = proteins significant in both analyses; orange = significant only in TAK vs HC; teal = significant only in LV-GCA vs HC; grey = non-significant in both analyses. E) Violin plots showing scaled protein levels (as Z-scores) for example proteins with prominent changes in both TAK and LV-GCA. P_adj_ < 0.05: *, P ≤ 0.01: **, P ≤ 0.001: ***, P ≤ 0.0001: ****.

As expected, many upregulated proteins in TAK and LV-GCA indicated immune activation including cytokines, chemokines, and growth factors (**Figure S2B**). Plasma IL6, the pleiotropic cytokine of known importance to LVV pathogenesis^13^, was significantly upregulated in both TAK and LV-GCA (**Figure 2E**), together with liver-derived inflammatory proteins such as SAA4 and FCN2. Prominent innate immune involvement in both TAK and LV-GCA was indicated by increased levels of neutrophil-derived proteins (S100A12, LCN2, DEFA1) and monocyte/macrophage activation and chemotactic factors (CSF1, CCL3, CCL5, CCL7, CCL14). Plasma levels of TNF (tumour necrosis factor), IL12B and IFNG were not significantly altered despite previous links to TAK pathogenesis^14^. In contrast, we observed large increases in OSM (oncostatin M) and TNFSF14 (LIGHT), cytokines not previously associated with LVV (**Figure 2E**). In addition to the dysregulation of immune-related proteins, we observed the upregulation of proteins with functions related to the extracellular matrix (TIMP1, MMP1, CST3, TNC), fibrosis (TGFB1) and angiogenesis (VEGFA, HGF, ANG, COL18A1) in both diseases, likely reflecting a signature of arterial injury and remodelling. Six proteins were consistently downregulated in both diseases, including IL7R, KIT, TNXB and THBS4 (both extracellular matrix (ECM) related glycoproteins), DPP4 (a glucose metabolism and T-cell activation factor) and CR2 (the complement C3d receptor).

To evaluate whether there were disease-specific effects, we performed a direct comparison of TAK versus LV-GCA, which revealed 6 significant DAP (**Table S5**). Visualising the relative abundance of these proteins in each group demonstrated that the dysregulation of these proteins appeared to be LV-GCA specific (**Figure S3A**). For example, in LV-GCA, but not TAK, CXCL9, CCL11 and CA3 levels were elevated compared to HCs. Similarly, CR2 and TNFSF11 (or RANKL) were significantly reduced in LV-GCA but not in TAK. Changes specific to TAK were less prominent, with no proteins that had significant changes in TAK versus both HCs and LV-GCA. However, we observed that FGF23 and IL17A were significantly increased in TAK compared to HCs but were not significantly increased in LV-GCA versus HC (**Figure S3B**).

### Signatures of active disease in TAK and LV-GCA patients

To identify proteins associated with disease activity in TAK and LV-GCA, we next compared the proteomic profile of active and inactive patients within each disease. In TAK, 16 proteins were significantly altered in active disease, with 11 upregulated and 5 downregulated (**Figure 3A**, **Table S6**). Upregulated proteins included neutrophil-related factors (S100A12, CXCL5 CXCL1), liver-derived proteins (SAA4, CFHR5), ECM components (TNC, NID1, CRTAC1, COMP) and angiogenic factors (VEGFA, ANG), indicating innate immune activation and vascular remodelling (**Figure 3B**). To corroborate the results of the active versus inactive patient analysis and identify proteins whose levels vary with the degree of disease activity, we tested for association with the numerical ITAS2010 disease activity score^15^. 8 of the 158 proteins measured were significantly correlated (adjusted P<0.05) with the ITAS2010 score (**Figure S4A & C**), and 7 of these 8 were differentially abundant in the active versus inactive analysis.

**Figure 3.**
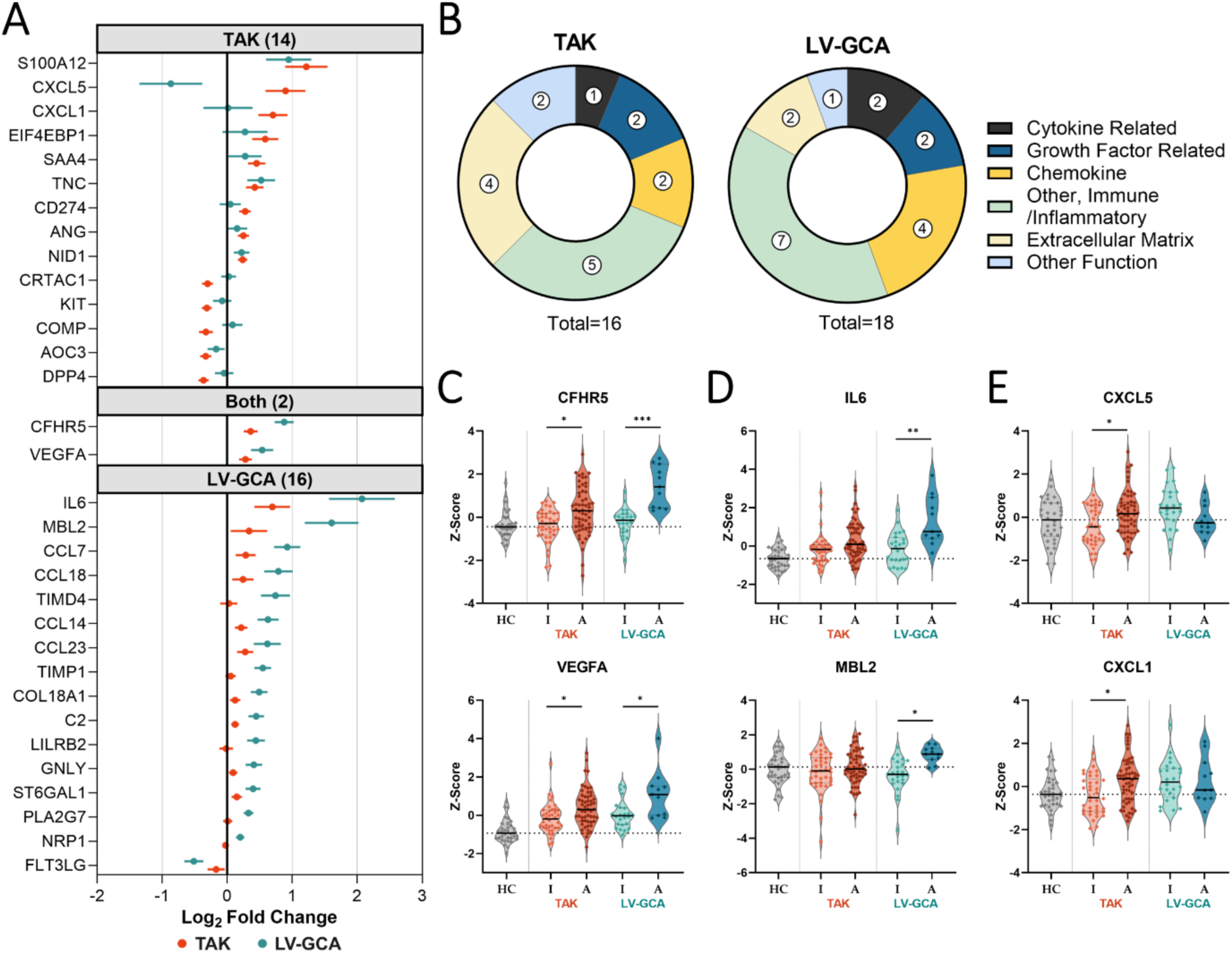
Proteins associated with active disease within TAK and LV-GCA patients. A) Points indicating log_2_ fold changes and standard error (horizontal bars) for proteins that were significantly (adjusted P < 0.05) differentially abundant in the active vs inactive patient analysis for Takayasu Arteritis only (TAK, N=56 vs N=40) (top panel), both diseases (middle panel) and Large Vessel Giant Cell Arteritis only (LV-GCA, N=11 vs N=24) (bottom panel). B) Functional categories of differentially abundant proteins from TAK and LV-GCA activity analyses. Violin plots depict the scaled abundance values (Z-scores) for proteins associated with disease activity in both diseases (C) and for the proteins which had divergent associations with disease activity in TAK and LV-GCA (D & E). P-values adjusted using Benjamini-Hochberg method; No symbol: non-significant, adjusted P < 0.05: *, P ≤ 0.01: **, P ≤ 0.001: ***.

Assessment of disease activity in TAK is currently based on the evaluation of clinical features, imaging, and clinical laboratory measures of inflammation, particularly CRP levels. However, CRP lacks both sensitivity and specificity for active TAK^7^. We found that 6 proteins (NID1, TNC, S100A12, CD274 (sPD-L1), DEFA1 and DPP4) were more strongly correlated with ITAS2010 than CRP. In addition, for the 20 active TAK patients who had normal CRP levels (<5mg/L), 12 (60%) had abnormal levels (defined as greater than the mean + 3 standard deviations of HCs) in one or more of the ITAS2010-associated proteins, indicating their potential added value as markers of disease activity (**Figure S4D**).

In LV-GCA, 18 proteins were significantly associated with active disease, with 17 increased and 1 decreased (**Figure 3A**, **Table S7**). Although two proteins (CFHR5 and VEGFA) were increased in active disease in both LV-GCA and TAK, the LV-GCA activity signature was largely distinct to that of TAK (**Figure 3A**). A more prominent acute phase response was evident in active LV-GCA compared to TAK with large increases in IL6 and multiple liver-derived inflammatory proteins (MBL2, ST6GAL1, C2) (**Figure 3D**). There were also differences in the chemokines and other immunoregulatory proteins affected; CXCL5 and CXCL1 levels were not significantly changed in active LV-GCA (**Figure 3E**) but there were increases in the monocyte-attracting chemokines (CCL7, CCL14, CCL23) and the T-cell recruitment and activation factors (CCL18, GNLY, TIMD4). Lastly, there were activity-associated increases in remodelling associated proteins TIMP1, COL18A1 and NRP1 in LV-GCA but not TAK.

To determine whether proteomic differences persist despite clinical remission, we compared inactive TAK and LV-GCA patients to HC participants. This analysis identified 22 and 61 DAP in inactive TAK and LV-GCA, respectively, with 18 common to both diseases (**Figure S5A-C**, **Tables S8 & S9**). Examples of proteins which remained elevated in inactive disease include OSM, S100A12, TNFSF14 and AXIN1 (**Figure S5D**). Importantly, these proteins remained chronically elevated regardless of disease duration (**Figure S5E**), even in TAK patients who had withdrawn all treatment following durable remission (**Figure S5D**; median time off treatment 2 years [inter-quartile range, IQR: 1.5-5.1]). Thus, our data indicate that a proportion of the proteomic changes observed in TAK and LV-GCA patients represent persistent molecular derangements which do not normalise with clinical remission.

### Biopsy proven C-GCA has a distinct proteomic endotype

We next performed proteomic profiling of an independent cohort (Cohort 2) of 239 patients presenting acutely with possible C-GCA, recruited to the TABUL study^16^. Blood samples were taken rapidly following initiation of high dose glucocorticoids; median treatment duration was 2 [IQR: 1-4] days. All patients underwent both temporal artery biopsy (TAB) and ultrasound sonography (USS) and a diagnosis of C-GCA was subsequently confirmed or excluded (Not C-GCA). Patient characteristics were typical for suspected C-GCA, with the majority of cases being over 60 years old (87.4%) and a predominance towards female sex and White European ancestry (**Table 2**). 56 (37.3%) of patients diagnosed with C-GCA had a positive TAB, 78 (52%) had an abnormal USS and 53 (35.3%) were negative for both TAB and USS with the diagnosis made on the basis of clinical features. Compared to Not C-GCA patients, C-GCA patients were slightly older (median age 4 years greater) and had higher ESR, CRP and platelet levels (**Table 2**). Proteomic profiling was performed using the same Olink platform. 167 proteins (91%) passed QC parameters and data from a minimum of 225 patients were available for analysis (**Methods**).

**Table 2:** C-GCA cohort.

|  | Not C-GCA | C-GCA |
| --- | --- | --- |
| N | 89 | 150 |
| Age | 69 [62, 76] | 73.5 [67, 78] |
| Female (%) | 63 (70.8) | 108 (72) |
| White European (%) | 88 (98.9) | 149 (99.3) |
| TAB+ (%)<sup>*</sup> | 0 (0) | 56 (37.3) |
| USS Positive (%) | 27 (30.3) | 78 (52) |
| Cranial Ischaemic Complications (%) | 18 (20.2) | 33 (22) |
| Polymyalgic symptoms (%) | 7 (7.9) | 18 (12) |
| ESR, mm/hr<sup>†</sup> | 13 [5, 28] | 35 [19.5, 58] |
| CRP, mg/L<sup>†</sup> | 14 [4, 22.5] | 30 [12.7, 54] |
| Platelets, 10<sup>9</sup>/L<sup>†</sup> | 290 [236, 332] | 348 [277, 454] |

*Data is median [IQR] or N (%) where indicated. <sup>\*</sup>Six patients did not have TAB result available. <sup>†</sup>Only* *partial data available: Not C-GCA N = 81, 55, 89 for ESR, CRP and platelets respectively; C-GCA N =* *143, 98, 147. TAB, temporal artery biopsy; USS, temporal artery ultrasound sonography; Cranial* *Ischaemic Complications (as per methods); ESR, erythrocyte sedimentation rate; CRP, C-reactive* *protein.*

Proteomic comparison of C-GCA patients (n=150) to Not C-GCA cases (n=89) revealed 31 DAP (**Figure 4A**, **Table S10**). Further investigation revealed heterogeneity of these protein profiles within C-GCA patients and that differences compared to Not C-GCA cases were mostly driven by the TAB positive (TAB+) C-GCA patient subset (**Supplementary Methods**, **Figure S6**). In analyses stratified by TAB result, comparison of TAB+ C-GCA (n=56) to Not C-GCA identified 62 DAP (**Figure 4B**, **Table S11**), while only 1 DAP was identified in the TAB negative (TAB-) C-GCA (n=89) versus Not C-GCA comparison (**Table S12**). The increase in significant associations when limiting to biopsy-proven cases despite reduction in sample size and hence statistical power indicates that TAB+ C-GCA is enriched for proteomic signal, and that the TAB-group were diluting this signal in the analysis of all C-CGA versus Not C-GCA. Comparison of the estimated log_2_ fold changes (log_2_FC) and protein abundances from the TAB+ and TAB-stratified analyses confirmed larger magnitudes of effect in the former (**Figure 4D & 4F**). We additionally found that TAB+ C-GCA patients had higher levels of CRP, ESR, platelets and presence of polymyalgic symptoms compared to TAB-C-GCA patients (**Figure 4C & Table S13**). Moreover, PCA of all proteins assayed indicated separation of TAB+ from TAB-C-GCA patients (**Figure S6C**). Together, these findings indicate that C-GCA can be stratified into biologically and clinically distinct subsets by TAB result.

**Figure 4.**
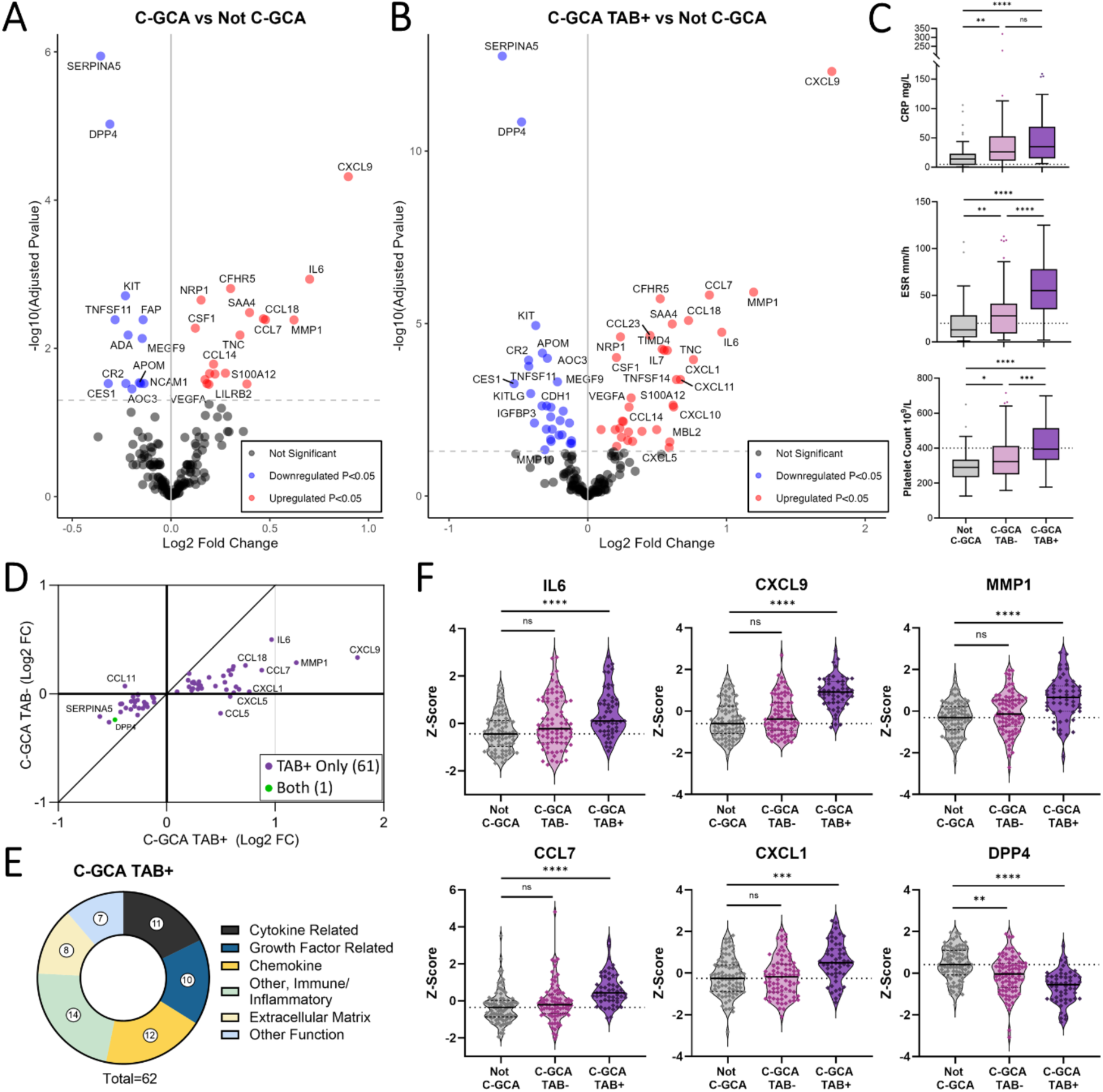
Proteomic changes associated with C-GCA are most pronounced in biopsy proven disease. A) Volcano plots for the differential protein abundance comparisons of Cranial Giant Cell Arteritis (C-GCA, N=150) cases vs Not C-GCA cases (N=89) and B) for the comparison of temporal artery biopsy positive C-GCA (C-GCA TAB+, N=56) versus Not C-GCA (N=89). −Log_10_(P_adj_) = −log_10_ Benjamini-Hochberg adjusted p-value. Red and blue indicate proteins that are significantly (P_adj_ < 0.05) upregulated and downregulated, respectively. C) Boxplot showing comparison of C-reactive protein (CRP), erythrocyte sedimentation rate (ESR) and platelet count between Not C-GCA, C-GCA TAB+ and C-GCA TAB-patients. Median and IQR represented by line and box edges respectively, upper whisker represents the upper quartile plus 1.5 times the IQR, lower whisker represents lower quartile minus 1.5 times IQR. Statistical comparisons made with Kruskal-Wallis and Dunn’s post-hoc tests. D) Comparison of log_2_ fold changes for differentially abundant proteins in C-GCA TAB+ vs Not C-CGA and C-GCA TAB- vs Not C-GCA. E) Functional categories of differentially abundant proteins in C-GCA TAB+ vs Not C-GCA comparison. F) Violin plots showing scaled protein levels (Z-score) for example proteins with prominent changes in C-GCA TAB+ cases. NS: non-significant, adjusted P < 0.05: *, P ≤ 0.0001: ****.

Further exploration of TAB-patients with hierarchical clustering revealed that while the large majority did not share the TAB+ associated signature, 18 TAB-patients were similar to TAB+ patients (**Figure S7**). However, this pattern had no significant association with demographic or clinical parameters. Relatedly, the comparison of C-GCA TAB-patients who had abnormal USS (N=36) to Not C-GCA cases did not identify any significant proteins. Together, these results suggest that TAB rather than USS positivity more closely reflects the proteomic phenotype.

The 62 proteins associated with TAB+ C-GCA patients suggest both innate and adaptive immune activation with dysregulation of cytokines, growth factors, chemokines and other immune-related proteins (**Figure 4E**). In a previous proteomic study^17^, five of these proteins were identified as significantly altered in C-GCA (**Figure S8** & **Table S11**). Similar to TAK and LV-GCA, many proteins involved in innate immune function were upregulated including acute phase response mediators (IL6, MBL2, SAA4, CFHR5, ST6GAL1) and factors involved in neutrophil and monocyte migration and activation (CXCL1, CCL14, CCL7, S100A12, CSF1). Several chemokines involved in T- and B-cell recruitment were also increased (CXCL9, CXCL10, CXCL11, CCL18) together with altered levels of lymphocyte survival and proliferation factors (increased IL7, decreased: IL7R, KITLG, TNFSF11). In addition, increases in ECM (MMP1, TIMP1, TNC, LTBP2), fibrosis (TGFB1) and angiogenesis-related proteins (VEGFA, HGF, NRP1) were indicative of vascular remodelling. The downregulated proteins with the lowest p-values were SERPINA5 and DPP4 (**Figure 4F**). In secondary analyses, we did not identify any proteins that were significantly associated with cranial ischaemic complications or polymyalgic symptoms within C-GCA patients when analysed both as a single group and when separated by TAB result.

### Correlated proteomic changes in active TAK, LV-GCA and biopsy-proven C-GCA with IL6 and VEGFA identified as key hub proteins

We next explored similarities and differences in the plasma proteomic signatures associated with each form of LVV. We considered the possibility that our results might be impacted by differences in study design. In Cohort 2, C-GCA patients were sampled with active disease at the time of diagnosis, whereas in Cohort 1, patients were sampled during both active and inactive disease over a range of disease durations. To mitigate against this, we re-analysed Cohort 1 restricting case samples to active disease only. These analyses revealed 68 and 69 differentially abundant proteins for active TAK versus HCs and active LV-GCA versus HCs respectively, of which the majority had also been significantly altered in the corresponding previous analyses using all cases (80.9% and 81.6% respectively, **Tables S14 & S15**). We then compared the results of these TAK and LV-GCA analyses to the proteomic associations identified in the TAB+ C-GCA versus Not C-GCA comparison in Cohort 2.

112 DAP were identified in one or more LVV type (subsequently referred to as LVV-associated proteins, **Figure 5A, Table S16**). Directional changes were highly similar with 74 (66.1%) having concordant changes (**Figure 5B**) and significant correlation between both TAK and LV-GCA profiles with that of TAB+ C-GCA (Pearson *r* 0.49 and *r* 0.69, respectively, both P<0.0001). Twenty-six proteins (23.2%) were dysregulated in all three diseases and 33 proteins (29.5%) were dysregulated in two. Of the 26 shared DAP, all had directionally concordant changes, including 20 upregulated and 6 downregulated proteins. We define these 26 proteins as the ‘pan-LVV signature’ (**Table S16**). Upregulated proteins included IL6, acute phase proteins (SAA4, CFHR5, ST6GAL1), monocyte and neutrophil factors (S100A12, CSF1), monocyte and lymphocyte chemokines (CCL5, CCL7, CCL3, CCL18, CCL23) and TNFSF14. Also increased were proteins related to arterial remodelling including VEGFA, HGF, MMP1, TIMP1 and the ECM glycoprotein Tenascin C (TNC). Decreases included IL7R, KIT and KITLG, each involved in lymphocyte differentiation and proliferation, DPP4, CR2 and TNXB (another ECM tenascin).

**Figure 5.**
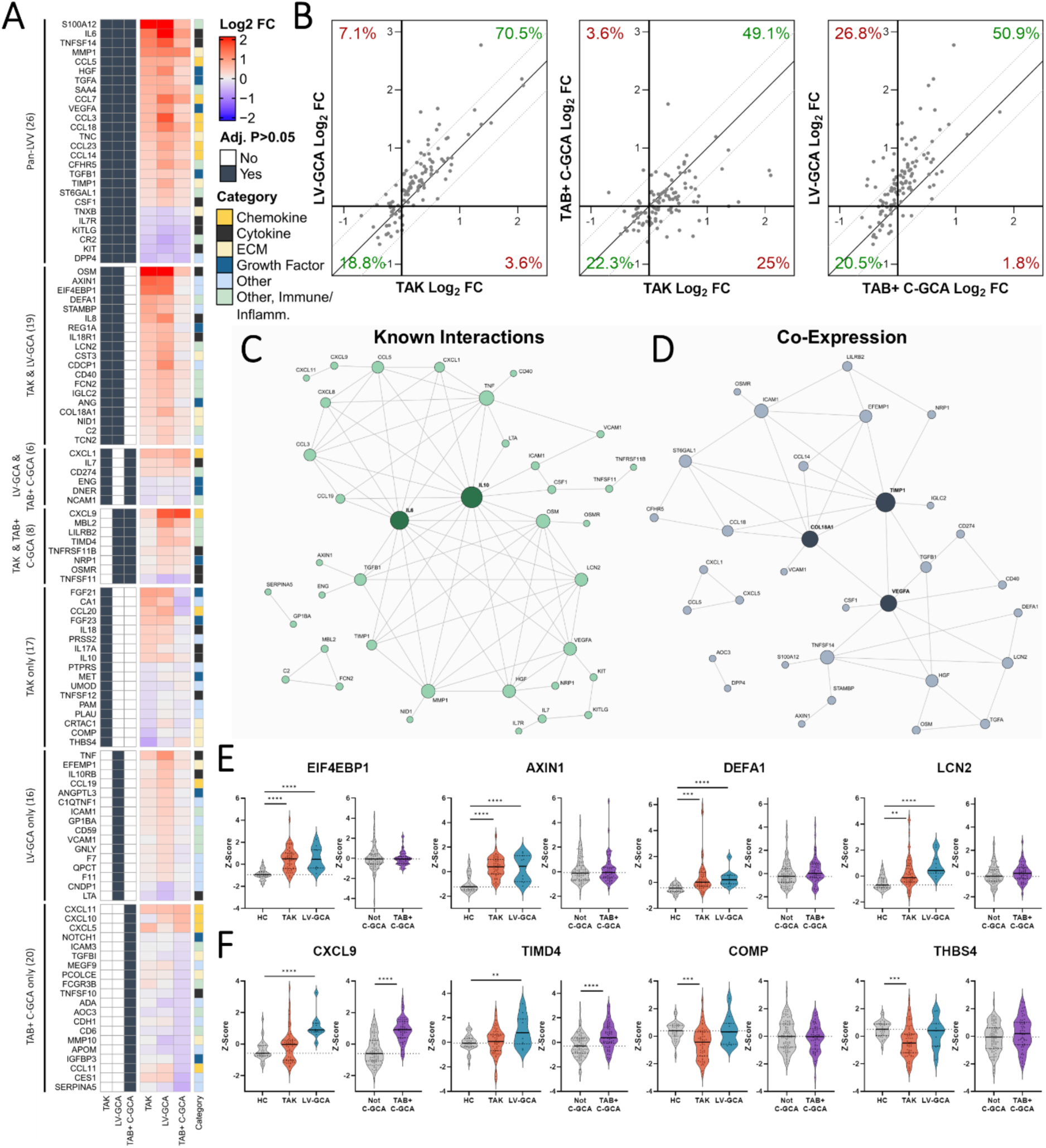
Comparison of active TAK, LV-GCA and biopsy-proven C-GCA proteomic profiles. A) Heatmap showing log_2_ fold changes (FC) of the 112 differentially abundant proteins (DAP) identified in active Takayasu Arteritis (TAK) vs healthy control (HC), active Large Vessel Giant Cell Arteritis (LV-GCA) vs HC and temporal artery biopsy positive (TAB+) C-GCA vs Not C-GCA comparisons. Left: navy boxes represent proteins with statistically significant changes (Adjusted P < 0.05) in each disease. Right: annotated functional category for each protein. B) Disease to disease comparison of Log_2_ FC for 112 DAP, diagonal lines represent the line of identity. Network plots of (C), known protein-protein interactions and (D), protein-protein co-expression for proteins with concordant changes in each disease (74). Node size corresponds to the number of connections to other nodes. For C, high confidence interactions (≥0.9) were sourced from STRING^20^ and for D, protein co-expression was defined as a Pearson correlation ≥0.6 in both active TAK/LV-GCA patients and TAB+ C-GCA patients. E) Violin plots showing scaled abundance values (Z-scores) for selected proteins associated with active TAK and LV-GCA but not C-GCA (F) and those identified as different between active LV-GCA and C-GCA vs active TAK. P-values adjusted using Benjamini-Hochberg method; No symbol: non-significant, Adjusted P < 0.05: *, P ≤ 0.01: **, P ≤ 0.001: ***, P ≤ 0.0001: ****.

Using the GTEx tissue transcriptome database^18^, we explored the global tissue expression profile of each LVV-associated protein, defining enhanced expression as >4 fold higher than average tissue level as per Human Protein Atlas methodology^19^. 83 (74.1%) had enhanced expression in ≥1 tissue. The most represented tissues were liver, spleen and whole blood (**Figure S9**). 11 proteins had enhanced arterial expression including TNC, TIMP1, COL18A1 and TNFRSF11B (OPG), thereby indicating the possibility of blood-based measurement of arterial biomarkers in LVV.

To infer relationships between proteins, we constructed two networks using the 74 proteins with concordant changes using i) annotated protein-protein interactions using String-db^20^ (**Figure 5C**) and ii) co-expression (Pearson *r*≥0.6) (**Figure 5D**). The network of annotated interactions identified IL6 and IL10 as the central hubs with connections to proteins with distinct functions including chemotaxis (e.g. CCL5, CCL3), angiogenesis (e.g. VEGFA, HGF) and tissue remodelling (e.g. MMP1, TIMP1, TGFB1). In the network constructed using protein co-expression data, TIMP1 and VEGFA displayed even greater connectivity, appearing as central nodes with edges connecting both to other remodelling-related proteins (e.g. COL18A1, NRP1) and multiple immune-associated proteins (e.g. CSF1, TNFSF14, CCL14). These networks indicate the coordinated regulation of immune and vascular remodelling processes, suggesting immune-stromal cross-talk in LVV.

The most prominent inter-disease differences diseases were between TAK and LV-GCA profiles compared to TAB+ C-GCA (**Figure 5A**). In particular, the large increases in EIF4EBP1 and AXIN1 observed in TAK and LV-GCA were not seen in C-GCA. Similarly, increases in the neutrophil proteins DEFA1 and LCN2 were observed exclusively in TAK and LV-GCA (**Figure 5E**). There were also differences between LV-GCA and TAB+ C-GCA profiles compared to TAK which indicate some degree of divergence between diseases. For example, increases in CXCL9 and TIMD4 were seen only in the GCA groups while decreases in the ECM-related proteins COMP and THBS4 were only found in TAK (**Figure 5F**).

### Plasma proteomic signatures reflect LVV arterial tissue phenotype

Plasma proteins arise not only from blood cells but also from a wide range of tissues and organs, including the vasculature which is in direct contact with the blood. We therefore sought to evaluate whether changes in patient plasma (**Figure 5A**) reflect the phenotype of arterial tissue affected by LVV. Using bulk RNA-seq data from a study which compared surgically resected aortic tissue of LV-GCA to non-inflammatory aortic aneurysms (NI-AA)^21^, we found that for 47 (42%) of the LVV-associated plasma proteins, the corresponding gene was differentially expressed in LVV arterial tissue (**Figure 6A, Table S17**). Moreover, the log_2_FCs of these 47 plasma proteins correlated with the log_2_FC of the corresponding gene in LVV tissue versus NI-AA (**Figure 6B**), and 28 (59.6%) had directionally concordant changes across the transcriptomic analysis of LVV arterial tissue and the plasma proteomic analyses of all the 3 LVV types.

**Figure 6.**
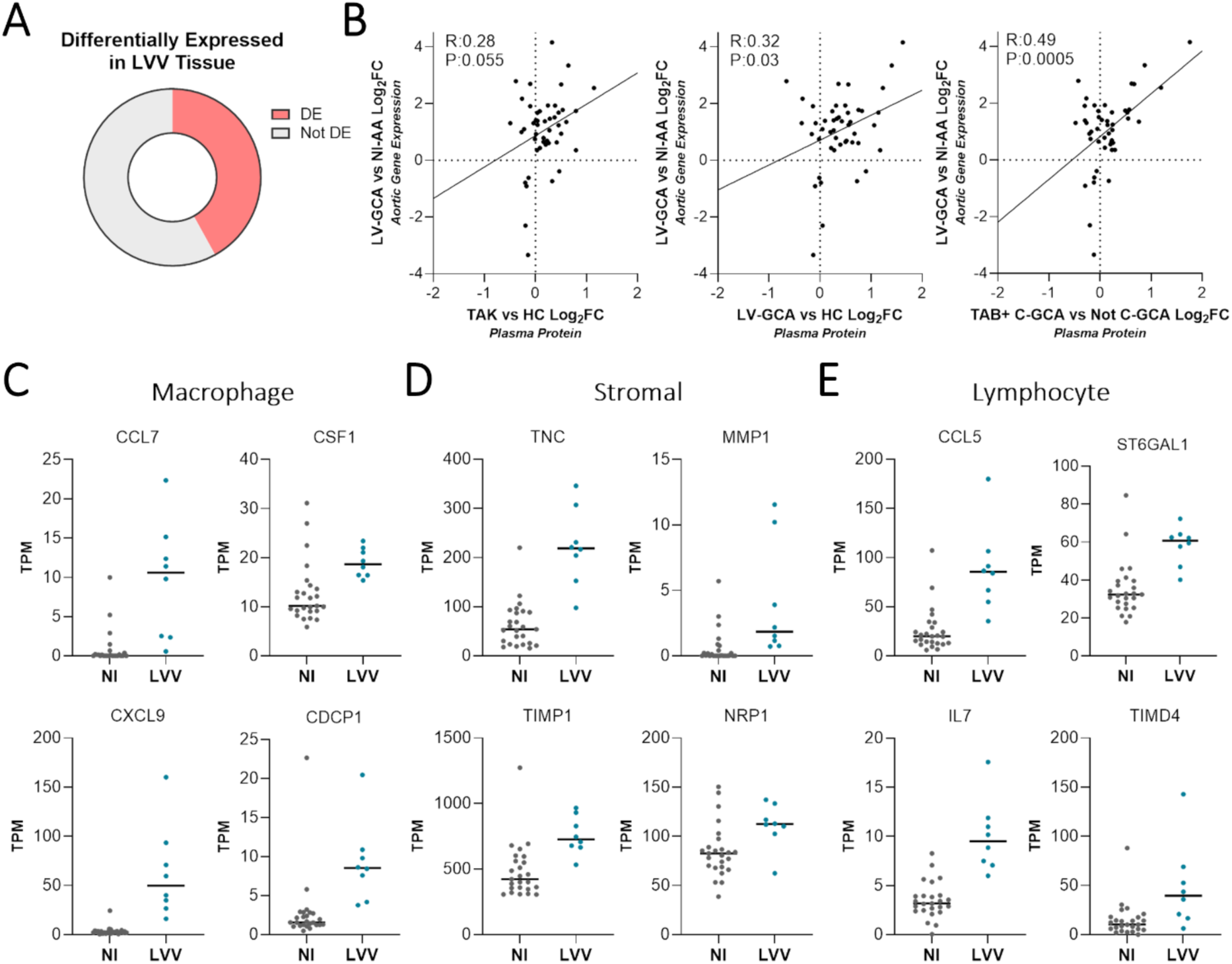
Correspondence of LVV plasma profile to arterial tissue phenotype. Comparison of plasma proteomic profiles associated with LVV and gene expression changes identified in aortic tissue affected by large vessel vasculitis (LVV)^21^. A) Pie chart showing the proportion of 112 LVV-associated plasma proteins (Table S16) that were also differentially expressed (Adjusted P < 0.05) in the comparison of large vessel giant cell arteritis (LV-GCA) related aortic aneurysm (N=8) to non-inflammatory aortic aneurysm (NI-AA) by bulk RNA-seq (N=25). B) Comparison of plasma protein and aortic gene expression log_2_ fold changes (Log_2_FC) for the 47 proteins and corresponding genes that had significant changes in both LVV plasma and aortic tissue, each point represents a gene/protein pair. For proteomics, Log_2_FCs represent active Takayasu Arteritis (TAK) vs healthy control (HC), active LV-GCA vs HC and temporal artery biopsy positive cranial-GCA (TAB+ C-GCA) vs Not C-GCA comparisons. For transcriptomics, Log_2_FCs represent LV-GCA associated aortitis vs NI-AA. (C-E), Dot plots showing aortic gene expression of genes/proteins DA in both LVV plasma and aortic tissue. Genes proteins typically expressed by macrophages (C), non-haematopoietic stromal cells (D) and lymphocytes (E). Gene expression measured in transcripts per million (TPM), cell-type expression classified using blueprint data (Figure S8). P-values were adjusted using Benjamini-Hochberg method.

Using the blueprint and GTEx bulk RNA-seq datasets^18,22^, we explored the cell-type and arterial expression profile of the 47 genes/proteins dysregulated in both plasma and arterial tissue (**Figure S10A**). Macrophage and neutrophil expressed genes/proteins made up the largest subset (34%); this included CCL7, CSF1, CXCL9 and CDCP1 which were increased in both tissue and plasma (**Figure 6C**). Genes/proteins expressed by non-immune stromal cells such as TNC, MMP1, TIMP1 and NRP1 were also prominent (**Figure 6D**). This latter cluster was enriched for genes of high arterial expression (GTEx) and could be useful as markers of arterial remodelling. The remainder were lymphocyte-derived (**Figure 6E**) or had mixed expression profiles. Of note, despite the significant decreases in plasma protein levels of DPP4, CR2 and IL7R in LVV, expression of the corresponding genes was significantly increased in LVV arterial tissue (**Figure S10B**), emphasising that there may be discordance of direction of effect in different tissue compartments. Overall, these findings indicate that plasma proteomic signatures can reflect aspects of LVV tissue phenotype and could provide a valuable non-invasive read-out of pathogenic processes occurring in diseased arteries.

## Discussion

TAK and GCA are currently classified as separate diseases^23,24^ but some investigators have proposed that they could represent varying manifestations of the same disease spectrum^4,5^. This debate has largely focused on phenotypic similarities, reflecting patterns of arterial injury^3^. Genome-wide association studies reveal differences in the genetic risk factors that predispose to TAK or GCA^25^. However, a key unanswered question is whether the molecular effector pathways acting in these diseases are shared or distinct. This information is critical for the rational selection of new therapeutic strategies that target specific proteins.

Here, we address this by comparing the plasma proteomic profile of TAK, LV-GCA and C-GCA. In 281 patients with LVV, we measured 184 inflammation- and vascular-associated proteins to characterise the plasma proteome of each major LVV type and evaluate changes associated with disease activity states. We found that the proteomic profiles associated with active TAK, LV-GCA and biopsy-proven C-GCA were similar and identified a 26-protein ‘pan-LVV’ signature common to all three groups. This signature primarily included proteins of immunological function, but it also comprised proteins arising from or acting on the stroma, indicative of arterial injury and/or repair. Some of these proteins have well-established roles in LVV (e.g. IL6 and VEGFA)^13,14^, but others have not been previously linked to LVV.

The signature reflected prominent innate immune activation, particularly with increases in several proteins related to monocyte and macrophage function. Importantly, co-expression analysis revealed coordinated regulation of such proteins (e.g. CSF1, CCL18, CCL14) with multiple proteins involved in tissue remodelling (e.g. VEGFA, TIMP1 and TGFB1) thereby highlighting innate immune-stromal crosstalk in LVV. This is consistent with recent reports implicating pro-fibrotic macrophage subsets in the fibrotic and stenotic remodelling of arteries in TAK and GCA, cell types that are less affected by current treatments^26,27^. These findings underscore the importance of macrophages in TAK and GCA pathogenesis and illustrate the need for markers and therapeutics which target macrophages beyond pro-inflammatory functions alone.

We also observed evidence for adaptive immune involvement, with changes in lymphocyte-related proteins like IL7R and TNFSF14. Plasma IL7R was decreased in all LVV groups, and its ligand IL7 was significantly increased in TAK and C-GCA. IL7 is essential for T-cell development and homeostasis whilst soluble (sIL7R) potentiates IL7 activity by enhancing bioavailability ^28^. The pattern observed is similar to findings in ANCA-associated vasculitis and tuberculosis infection but contrasts with rheumatoid arthritis and lupus where sIL7R levels are increased^29–31^. As another example, TNFSF14 (LIGHT) was prominently elevated. TNFSF14 acts as a T-cell costimulatory factor and triggers T-cell activation and proliferation^32^. TNFSF14 promotes systemic immunopathology, as demonstrated by transgenic animals with constitutive TNFSF14 expression in T-cells^33^. Moreover, TNFSF14 can also act on non-haematopoietic structural cells including fibroblasts, endothelial and smooth muscle cells to drive tissue fibrosis^34^. Consistent with this, our co-expression network analysis identified an edge connecting TNFSF14 to VEGFA, a central hub node. Current treatment strategies in LVV are limited to suppression of inflammation and do not target fibrosis directly. Thus, TNSF14 antagonism may be a novel therapeutic approach in LVV that provides dual targeting of both the immune response and the consequent stromal reaction.

Lastly, the signature included tissue remodelling proteins expressed by immune cells (e.g. VEGFA) or stromal cells (e.g. TNC, TIMP1, MMP1); the latter may represent useful markers of arterial damage independent of inflammation. Increased plasma TNC in LVV is relevant given its enrichment in normal arteries and increased expression in arteries affected by LVV. TNC is an ECM protein primarily expressed by fibroblasts at sites of tissue damage where it supports repair. It gained interest as a candidate marker of vascular injury, but its levels are also elevated in other disease states^35^. Although its lack of specificity may preclude its use as a diagnostic marker, TNC and other ECM proteins could be useful for monitoring arterial injury during follow-up and should be evaluated in longitudinal studies.

Despite the overall similarity in LVV proteomic profiles, we identified differences which suggest some immunological divergence between diseases. For example, the chemokine CXCL9 was higher in LV-GCA and C-GCA compared to TAK. CXCL9 is produced by macrophages and other cell types in response to interferon-gamma and typically reflects activation of the Th1 pathway^36^. Increased plasma CXCL9 has been shown previously in GCA and is associated with CXCR3+ cell infiltration into diseased arteries^37^. Previous studies speculated that TAK and GCA differ in the susceptibility of T-cell pathways to glucocorticoids whereby Th1 responses persist in GCA and Th17 responses persist in TAK after treatment initiation^38,39^. Our observation that CXCL9 was exclusively increased in GCA while IL17A was exclusively increased in TAK may support this theory.

Beyond comparing LVV types, this study provided several disease-specific insights. We identified novel markers of active disease in TAK and in LV-GCA. Disease activity assessment is challenging in LVV, where inflammatory markers such as CRP may not correspond to disease activity detected in radiological or histopathological examination^7,40^. Further exploration within TAK patients identified six proteins more strongly associated with ITAS2010 score than CRP, and we show that their use would provide more sensitive detection of active disease. Importantly, we also found that many proteins remained altered in TAK and LV-GCA patients despite remission. OSM and AXIN1 were elevated regardless of activity, disease duration or treatment withdrawal. Activity markers like S100A12 and VEGFA were highest in active disease but remained increased in inactive disease. These findings likely reflect persistent low-grade inflammation and are analogous to the residual molecular signatures previously described in a multi-omic study of remission in rheumatoid arthritis^41^. However, whether persistent molecular abnormalities impact clinically important outcomes remains unknown.

Our results suggest that biopsy-positive and negative C-GCA are proteomically distinct. There are two possibilities that could explain our findings. First, despite a careful clinical phenotyping algorithm which included external expert review, there may be instances of misclassification within the TAB-C-GCA group, such that some patients were erroneously labelled as C-GCA. However, this cannot fully explain our findings since the TAB-C-GCA group included some patients with a positive USS. Alternatively, TAB positivity may reflect the burden of arterial disease. Skip lesions, with discontinuous segments of arterial inflammation, can occur in GCA and so a patient with lower burden of arterial disease is more likely to have a negative biopsy on the small biopsy section of artery sampled. Thus, it is possible that the proteomic read-out may reflect quantitative differences in the extent of arteritis. Our clinical lab data provided additional evidence of differences between TAB+ and TAB-patients, with higher ESR, CRP and platelet count in the former group, in keeping with previous studies^42–44^. Other clinical differences in TAB+ and TAB-C-GCA have been described, including greater risk of visual loss^44,45^ and higher prevalence of PMR in TAB+ patients^42^, although these findings have not replicated in all case series^46^. Our data support the concept that C-GCA can be stratified by biopsy into biologically distinct endotypes, which may have potential implications for future trial design and precision medicine strategies.

Our study had certain limitations. We used a targeted proteomic panel enriched for inflammatory and vascular proteins and thus our ability to compare between LVV groups is limited to the proteins measured. The HCs in Cohort 1 were well-matched to TAK patients but were younger than LV-GCA patients. Differences in study design between Cohort 1 and Cohort 2 mean that proteomic differences between C-GCA and the other LVV groups may be confounded by differences in disease duration and treatment history. In addition, the controls in Cohort 2 (“Not GCA”) were individuals who presented with symptoms for which a diagnosis of C-GCA was considered but ultimately excluded, whereas the controls in Cohort 1 were healthy participants with no symptoms. In Cohort 1, there was heterogeneity in treatment, with varying use of steroids and other immunosuppressants which could potentially impact the plasma proteome. However, treatment (particularly glucocorticoid dose) is given in response to disease activity, reflected in the correlation between disease activity scores and prednisolone dose in Cohort 1. Thus attempting to statistically adjust for treatment risks over-adjustment^47,48^. Finally, in the case of membrane-bound proteins that undergo cleavage to produce a soluble form, it is not always clear whether plasma protein measurements are exclusively capturing the latter or also protein from cell membranes (for example, arising from *in vivo* sources such as exo-/ectosomes or *ex vivo* processes such as venepuncture or sample processing), complicating interpretation.

In conclusion, similarities in the plasma proteomic profiles of active TAK and GCA indicate common effector pathways resulting in inflammatory arterial damage despite differences in genetic aetiology. Our integrated analysis of plasma and arterial tissue highlight the role played by macrophages and their protein products in LVV and indicate significant potential for their targeting in novel treatments and biomarkers. Future work should expand the molecular characterisation of the LVV disease spectrum by extending the number of proteins measured via use of complementary proteomic platforms and by concurrent measurement of other -omic layers (e.g. RNA-seq of immune cells). Longitudinal studies characterising the temporal changes in the molecular profile across the disease course will be valuable in delineating acute from chronic changes and allowing intra-individual assessment of putative biomarkers identified here.

## Methods

### Cohort 1 study participants

Patients with TAK or LV-GCA were recruited from the Hammersmith Hospital (Imperial College Healthcare NHS Trust, UK) between 2013 and 2020. TAK patients fulfilled EULAR/ACR 2022 classification criteria^23^ and all had typical patterns of arterial involvement in radiological assessments. LV-GCA patients were >50 years at onset with radiological evidence of LVV, as defined previously^13^. Three LV-GCA patients had concurrent temporal artery involvement (confirmed by temporal artery USS and/or TAB). HC participants were recruited locally from hospital and college staff and had no history of inflammatory or cardiovascular disease. Citrate blood samples were centrifuged at 1000G for 10 minutes within 4 hours of venepuncture and plasma was stored at −80°C until use. Disease activity was assessed using the Indian Takayasu Clinical Activity Score (ITAS2010) for TAK^15^ and the National Institutes of Health (NIH) score for LV-GCA^40^. Active disease was defined as ITAS2010 score ≥ 1 or ITAS-CRP ≥ 2 for TAK and NIH score ≥ 2 for LV-GCA. All inactive cases were retrospectively confirmed to be relapse free for 1 year following sample collection. Patients and HC provided written informed consent, and samples were collected as a sub-collection registered with the Imperial College Healthcare Tissue Bank (licence: 12275; National Research Ethics Service approval 17/WA/0161).

### Cohort 2 study participants

The <u>T</u>emporal <u>A</u>rtery <u>B</u>iopsy vs <u>UL</u>trasound in Diagnosis of GCA (TABUL) was an international, multicentre, prospective study which compared the sensitivity and specificity of temporal artery ultrasound to biopsy in 381 patients with suspected C-GCA^16^ [ClinicalTrials.gov: NCT00974883]. Reference diagnosis of C-GCA or Not C-GCA at 6 months was based on a combination of baseline signs and symptoms, blood tests, TAB, fulfilment of ACR 1990 GCA classification criteria, clinical course during the follow-up period, final consultant diagnoses and verification by an expert review panel, as described previously^16^.

Cranial ischaemic complications were defined as permanent ocular or non-ocular conditions at presentation. Ocular complications: anterior ischaemic optic neuropathy, branch retinal artery occlusion, cilioretinal artery occlusion, cranial nerve palsy (III, IV or V), central retinal artery occlusion, posterior ischaemic optic neuropathy relative afferent pupillary defect; irreversible visual loss; irreversible visual field defect; irreversible ocular motility or irreversible diplopia. Non-ocular cranial complications: scalp necrosis; tongue necrosis; cerebrovascular accident at presentation considered secondary to GCA). Polymyalgic symptoms at presentation are also reported but do not represent a confirmed diagnosis of polymyalgia rheumatica.

Citrated blood samples collected as soon as feasible after starting glucocorticoid treatment (median [IQR] 2 [1-4]) and were centrifuged at 2500 G for 15 minutes within 1.5 hours of collection and plasma was stored at −80°C until use. Due to funding constraints and biosample availability, we performed Olink proteomic assays on 239 patient samples out of the 381 patients recruited to TABUL. We included all available samples from patients with a diagnosis of C-GCA. We selected a subset of sex and age (+/- 5 years) matched Not C-GCA cases such that the ratio of C-GCA to Not C-GCA was approximately 2:1. As part of the study design, we selected an equal proportion of cases with cranial ischaemic complications in the C-GCA group as in the Not C-GCA group. Overall study approval was granted by the Berkshire Research Ethics Committee (09/H0505/132) and approved was also granted at local participating clinical sites.

### Proteomic analysis

184 proteins were measured by proximity extension assay using two Olink Target panels, ‘Inflammation 1’ and ‘Cardiometabolic’ at the Leeds Immunogenomics Facility, University of Leeds. In order to provide a succinct and standardised nomenclature, we report proteins by the symbols of the genes encoding them (see **Table S3** for a full list of proteins and associated full names and accession numbers). Cohort 1 and 2 samples were processed and analysed independently, but proteomic measurements were performed in the same facility. We designed assay plates such that samples were balanced across plates according to disease grouping and disease activity status, with randomisation to determine well position within plates. Proteomic data were normalised using standard Olink workflows, which includes inter-plate normalisation, to produce measures of relative protein abundance (‘NPX’) (log_2_ scale). For visualisation, we transformed to Z-scores (mean=0, SD=1). Due to a technical fault with a PCR machine during the running of one plate on the Inflammation 1 panel for Cohort 2, it was necessary to re-run this plate as a separate batch. Principal component analysis (PCA) revealed a batch effect, with samples from this plate separated from samples on the other three plates run as part of the first batch. We therefore adjusted for this batch effect for proteins measured on the Inflammation 1 panel in Cohort 2, using batch (a binary variable) as a covariate in linear model based differential abundance analyses. For situations requiring batch-correction outside of differential abundance testing (e.g. visualisation of protein levels using violin plots and heatmaps and network analyses), the residuals from the linear model (in Wilkinson notation) NPX ~ Batch were used to generate batch-corrected protein values. Further PCA of these residuals confirmed that the batch effect had been removed. The residuals were then converted to Z scores prior to visualisation or other downstream analyses. Cardiometabolic panels for Cohort 1 were not affected by this issue and were analysed as a single batch.

Proteins with >75% of samples below the lower limit of detection were removed resulting in 158 and 167 protein measurements for Cohort 1 and 2, respectively. Sample-level QC was performed using internal assay controls, boxplots of relative protein abundance values and PCA for outlier detection. In Cohort 1, three samples (1 HC and 2 TAK) were excluded from Inflammation panel measurements due to amplification failures. In Cohort 2, fifteen samples (9 C-GCA and 6 Not C-GCA) were excluded from Inflammation panel and 3 (all C-GCA) from the Cardiometabolic panel measurements due to amplification failures and flagged status in internal QC checks. Wherever possible all available data was analysed (e.g. differential abundance analyses) but some analyses (e.g. hierarchical clustering, PCA, multiple linear regression versus clinical parameters) necessitated using only complete data. The final post-QC datasets are provided on figshare: **<u>10.6084/m9.figshare.26928211</u>**.

Differential protein abundance was performed using linear models in R. For a given protein, protein abundance was regressed on disease status (encoded as 0 or 1). The beta coefficient for the disease status term represents the estimated log_2_ fold change in the protein level between groups under comparison. For example, for the analysis of TAK versus HCs, the regression model was NPX ~ D, where NPX was log_2_ protein level (continuous variable) and D was a binary variable, encoded as 0 for HC and 1 for TAK. Correction for multiple testing (multiple proteins) was performed using the Benjamini-Hochberg method and an adjusted P-value of < 0.05 (i.e. false discovery rate < 5%) was defined as significant.

### Protein annotation

Olink panel proteins were manually classified as “Cytokine Related”, “Growth Factor Related”, “Chemokine, Other Immune/Inflammatory related protein”, “Extracellular matrix Related” or “Other Function” for the purpose of the annotation of differential abundance results. This was done using a combination of public resources including Gene Ontology terms, pathway and functional databases. The full list of proteins and associated classifications is provided in **Table S3**.

### Network analysis

The protein-protein interaction network between differentially abundant proteins was constructed using high confidence interactions (confidence ≤ 0.9) sourced from STRING^20^. No additional filtering of interactions was performed. The protein co-expression network of differentially abundant proteins was created using inter-protein correlation. Node edges were defined as Pearson *r* ≥ 0.6. Cohort 1 (TAK and LV-GCA) and Cohort 2 (C-GCA) networks were computed individually and then intersected so that only correlations present in both networks feature in the final network. Both networks were plotted using the igraph package in R^49^.

### Tissue expression of differentially abundant proteins

GTEx bulk RNA-seq tissue expression data was accessed as median transcript per million values per tissue^18^. Data pre-processing included: removal of sex-specific organ data (i.e. cervix, breast, vagina, testis, fallopian tubes), removal of purified cell data (e.g. cultured fibroblasts) and where there were multiple sample types per tissue group (e.g. Artery-Coronary or Artery-Aorta) the highest expression value was used for that tissue type. Enhanced tissue expression of differentially abundant proteins was defined as >4 fold higher than the averages expression in other tissues as done previously by the Human Protein Atlas^19^.

### RNA-seq analysis of LVV arterial tissue

A previous study compared the transcriptomic profile of inflammatory and non-inflammatory aortic aneurysms using bulk RNA-seq^21^. Gene-level count data was accessed and filtered for cases of inflammatory aneurysm associated with GCA (n=8) for comparison with non-inflammatory cases (n=25). Data was normalised, genes with low expression were removed and groups were compared using the standard edgeR package methodology^50^. Differentially expressed (DE) genes were defined using Benjamini Hochberg adjusted P < 0.05. DE genes were then compared with LVV-associated plasma proteins with regards to overlap and Pearson correlation of log_2_ fold change in each disease.

### Other analyses

Details of additional analyses are provided in **Supplemental Information File**.

## Data availability

Individual-level normalized post-QC proteomic data are provided on figshare: **10.6084/m9.figshare.26928211**

## Acknowledgments

We thank Prof. Marina Botto and Prof. Matthew Pickering for helpful comments on the manuscript. We thank the patients and healthy volunteers who participated in this study, along with the clinical research teams who recruited patients. We thank Surjeet Singh as the chief study coordinator for TABUL, and Bhaskar Dasgupta as a co-investigator on TABUL who played a leading role in the study recruitment. Prof Mason tragically passed away during the study and we dedicate this paper to his memory.

## Supplemental Material

**<u>Supplementary Information File.docx</u>**

Supplementary Methods

Figure S1: Comparing TAK findings to those of a previous plasma proteomic study

Figure S2: TAK and LV-GCA vs HC comparison

Figure S3: Plasma protein differences between TAK and LV-GCA patients

Figure S4: Proteins associated with Disease Activity Score in TAK

Figure S5: Differentially abundant proteins in inactive TAK & LV-GCA

Figure S6: Proteomic changes in C-GCA are most pronounced in biopsy proven disease

Figure S7: Comparison of proteomic changes in TAB+ and TAB-C-GCA patients

Figure S8: Comparing biopsy proven C-GCA findings to those of a previous plasma proteomic study

Figure S9: Tissue expression of LVV-associated plasma proteins

Figure S10: Cell-type expression profile of proteins/genes identified as dysregulated in both LVV plasma and tissue

**<u>Supplementary Data File.xlsx</u>**

Table S1: Proteins measured in this study

Table S2: Characteristics of TAK and LV-GCA patients with inactive and active disease

Table S3: Differential protein abundance analysis TAK vs HC

Table S4: Differential protein abundance analysis LV-GCA vs HC

Table S5: Differential protein abundance analysis TAK vs LV-GCA

Table S6: Differential protein abundance analysis Active TAK vs Inactive TAK

Table S7: Differential protein abundance analysis Active LV-GCA vs Inactive LV-GCA

Table S8: Differential protein abundance analysis Inactive TAK vs HC

Table S9: Differential protein abundance analysis Inactive LV-GCA vs HC

Table S10: Differential protein abundance analysis C-GCA vs Not C-GCA

Table S11: Differential protein abundance analysis C-GCA TAB+ vs Not C-GCA

Table S12: Differential protein abundance analysis C-GCA TAB- vs Not C-GCA

Table S13: Characteristics of Not C-GCA cases and C-GCA patients stratified by TAB result

Table S14: Differential protein abundance analysis Active TAK vs HC

Table S15: Differential protein abundance analysis Active LV-GCA vs HC

Table S16: Comparison of active TAK, active LV-GCA and C-GCA TAB+ proteomic profiles

Table S17: Proteins & Genes dysregulated in both LVV plasma and arterial tissue

*Note, Tables S2 and S13 are also provided in the supplementary information file for convenience*.

